# Clinical Biomarker-Based Biological Aging and the Risk of Abdominal Aortic Aneurysm: A Prospective Cohort Study

**DOI:** 10.1101/2024.12.16.24319128

**Authors:** Xinyi Zhou, Zaixin Yu, Mukamengjiang Juaiti, Lihuang Zha, Baohua Peng, Chengshuang Xiao, Yiyang Tang

**Affiliations:** Department of Cardiology, Xiangya Hospital, Central South University, Changsha, Hunan, China; National Clinical Research Center for Geriatric Disorders, Xiangya Hospital, Central South University, Changsha, Hunan, China

**Keywords:** Biological aging, Abdominal aortic aneurysm, UK Biobank, Genetic susceptibility

## Abstract

**Background:** Compared to chronological age, biological age (BA), a concept introduced in recent years, more accurately reflects the true aging status of the body. While biological aging has been found to be associated with various cardiovascular diseases, its relationship with abdominal aortic aneurysm (AAA) remains unclear.

**Methods:** This study utilized data extracted from UK Biobank for analysis. Telomere length (TL), and BA acceleration calculated using the Klemera-Doubal (KDM) and PhenoAge methods, were employed as surrogate measures to assess biological aging. Cox regression was primarily performed to explore the association between biological aging and AAA risk. Genetic susceptibility was assessed by constructing a polygenic risk score (PRS).

**Results:** This study included 311,646 participants, predominantly women and White, with a median age of 58 years. During a median follow-up of 12.54 years, 1,339 new cases of AAA (4.3‰) were reported. Each standard deviation (SD) decrease in TL was associated with a 20% increased risk of AAA (HR=1.20, 95% CI=1.14-1.27); each SD increase in KDM-BA acceleration was associated with a 21% increased risk (HR=1.21, 95% CI=1.12-1.29); and each SD increase in PhenoAge acceleration was associated with a 40% increased risk (HR=1.40, 95% CI=1.32-1.48). These associations were independent of genetic risk, as assessed by the PRS, and demonstrated a joint effect with genetic predisposition on the long-term risk of AAA.

**Conclusion:** Accelerated biological aging was longitudinally associated with an increased risk of AAA, suggesting it may be a significant factor and potential biomarker for the occurrence of the condition.

## Background

Abdominal aortic aneurysm (AAA) is the most common form of aortic aneurysm, characterized by the permanent and localized dilation of the abdominal aorta[1]. It is often asymptomatic until rupture, a catastrophic event with a mortality rate of up to 80%, highlighting its significant public health burden[2]. Therefore, identifying and validating biomarkers associated with AAA occurrence is of significant public health importance, as it can enhance early diagnosis, improve management strategies, and ultimately optimize patient outcomes.

AAA is a degenerative disease associated with aging[3]. Traditionally, chronological age has been used to assess an individual’s level of aging and disease risk[4]. However, chronological age has limitations, as individuals of the same or similar chronological age can exhibit significant differences in functional status and aging processes due to factors such as genetics, environment, and lifestyle[5]. Recently, biological age has been proposed as a more comprehensive and accurate measure of aging, incorporating indicators such as telomere length (TL), Klemera-Doubal method biological age (KDM-BA), and phenotypic age (PhenoAge)[6]. Previous studies have shown that increased biological age is significantly associated with higher mortality and elevated risks of various cardiovascular diseases, including heart failure[7, 8]. However, the relationship between biological aging and the incidence of AAA remains unclear. Additionally, AAA has a genetic predisposition, with genome-wide association studies (GWAS) identifying several single-nucleotide polymorphisms (SNPs) linked to AAA risk[9, 10]. The interaction between genetic susceptibility and biological aging in relation to AAA risk has not yet been fully explored.

To fill this research gap, this study uses a prospective cohort from the UK Biobank to examine the longitudinal associations between biological aging and AAA risk, and to investigate the joint effects and interactions between genetic susceptibility and biological aging on the long-term risk of AAA.

## Methods

### Study population

The data used in this study were obtained from the UK Biobank (ID: 107175). The UK Biobank is a large-scale prospective cohort study that recruited over 500,000 volunteers across the United Kingdom between 2006 and 2010[11]. This study received ethical approval from the North West Multi-Centre Research Ethics Committee (Ref: 11/NW/0382), and all participants provided informed consent by signing a consent form.

Of the 502,163 participants in the UK Biobank, we excluded 460 participants with baseline AAA, 177,504 participants missing biomarkers required for the KDM-BA and PhenoAge algorithms, 11,531 participants missing TL measurements, and 1,022 participants with missing covariate data. A total of 311,646 participants were included in the final analysis. The participant selection process was shown in **Figure S1**.

### Study Outcomes

The outcome variable of this study was the occurrence of AAA, which was determined based on hospital admission and death registration records (**Table S1**). The diagnosis of AAA was identified using the Tenth Revision of the International Classification of Diseases (ICD-10 codes: I71.3, I71.4) and the Office of Population, Censuses and Surveys: Classification of Interventions and Procedures (OPCS-4 codes: L18*, L19*, L254, L27*, L28*, L464), as used in prior studies[12]. The study period began when participants entered the cohort and ended upon the diagnosis of AAA, loss to follow-up, death, or completion of follow-up, whichever occurred first. The follow-up cut-off date was September 30, 2021, for England and Wales, and October 31, 2021, for Scotland.

### Measurement of TL

TL was measured using a multiplex quantitative polymerase chain reaction method, with samples obtained from peripheral blood leukocytes. Specifically, TL was quantified relative to the ratio of telomere repeat copy number to single-copy gene (T/S), and the data were log-transformed and Z-standardized. Detailed information on the measurement and adjustment of TL can be found in previous studies[13].

### Calculation of KDM-BA and PhenoAge

The KDM-BA and PhenoAge algorithms[8, 14, 15] are widely recognized biological age measures based on clinical parameters (**Table S2**). These algorithms were developed using optimal training models derived from the National Health and Nutrition Examination Survey (NHANES) and have been further validated with data from the UK Biobank. Using the R package “BioAge” (https://github.com/dayoonkwon/BioAge), KDM-BA was calculated based on forced expiratory volume in one second, systolic blood pressure, and seven blood chemistry parameters: albumin, alkaline phosphatase, blood urea nitrogen, creatinine, C-reactive protein (CRP), glycated hemoglobin, and total cholesterol. PhenoAge was calculated using nine hematological indicators: albumin, alkaline phosphatase, creatinine, CRP, glucose, mean corpuscular volume, red cell distribution width, white blood cell count, and lymphocyte percentage. We then regressed the calculated biological age values against the chronological age at the time of biomarker measurement and computed the residuals[16]. These residuals were termed “age acceleration” (AA) values, which were used to assess biological aging. The AA values were then standardized with a mean of 0 and a standard deviation of 1 for comparison in subsequent analyses. More details can be found in **Method S1**.

### Covariates

The covariates considered in this study mainly included age, sex, ethnicity, education level, Townsend Deprivation Index (TDI), CRP, body mass index (BMI), smoking and alcohol consumption status, physical activity, sleep and dietary patterns, as well as medical history of hyperlipidemia, hypertension, and diabetes. Detailed definitions and descriptions of these covariates can be found in **Method S2**.

### Statistical analysis

The continuous variables were found not to follow a normal distribution based on the quantile-quantile plot and were therefore expressed as medians [interquartile range (IQR)]. Group comparisons were performed using the Mann-Whitney U test or Kruskal-Wallis H test. Categorical variables were represented as frequencies (percentages), and group comparisons were conducted using chi-square tests.

TL, KDM-BA, and PhenoAge acceleration were treated as continuous variables or categorized based on quartiles, and their associations with AAA occurrence were assessed using Cox proportional hazards models, with results presented as hazard ratios (HR) and 95% confidence intervals (95% CI). The Schoenfeld residual test confirmed no violations of the proportional hazards assumption. Model 1 adjusted for age, sex, ethnicity, education level, and TDI. Model 2 further adjusted for smoking and drinking status, physical activity, sleep and dietary patterns, BMI, CRP, and medical history of hypertension, hyperlipidemia, and diabetes. Additionally, based on Cox regression Model 2, a restricted cubic spline (RCS) with three knots was used to analyze the dose-response relationship between biological aging and AAA risk.

We included only participants of White ancestry and constructed a polygenic risk score (PRS) to assess individuals’ genetic susceptibility to AAA[17] (**Method S3**). We then evaluated the joint effect of genetic susceptibility and biological aging on the risk of AAA. Participants were categorized into 12 groups based on their levels of biological aging and genetic susceptibility. Using the group with the lowest levels of both biological aging and genetic susceptibility as a reference, we estimated the HR and 95% CI for AAA occurrence in the other groups, after adjusting for covariates in Model 2. Stratified analyses were conducted to assess the separate effect of biological aging on AAA incidence at different levels of genetic susceptibility, and the likelihood ratio test was used to evaluate the interaction between biological aging and genetic susceptibility.

Subgroup analyses were also conducted to assess potential modifying effects on the association between biological aging and the risk of AAA. Sensitivity analyses included: 1. Excluding participants who developed AAA within the first year of follow-up; 2. Using the Fine-Gray model to account for mortality as a competing risk. All analyses were performed in R (version 4.3.2), with *p*-values < 0.05 considered statistically significant.

## Results

### Baseline of the participants

This study included a total of 311,646 participants (**Figure S1**), of whom 168,249 (54.0%) were women. The majority of participants were White, comprising 294,574 individuals or 94.5% of the total cohort (**Table S3**). The median age was 58 years (IQR: 50–63 years), with median values of KDM-BA and PhenoAge recorded at 49.82 years (IQR: 40.90-58.26) and 45.83 years (IQR: 38.10-52.54), respectively. KDM-BA (r = 0.67) and PhenoAge (r = 0.86) were strongly positively correlated with chronological age, whereas TL showed a weak negative correlation (r = -0.20, **Figure S2**).

Baseline characteristics, stratified by quartiles of TL, PhenoAge acceleration, and KDM-BA acceleration, were summarized in **Table 1** and **Tables S4–S5**. Participants with shorter TL were more likely to be men, have lower educational attainment, and engage in unhealthy lifestyle behaviors (e.g., current smoking, poor sleep patterns, and insufficient physical activity). They also exhibited higher prevalence rates of diabetes, hyperlipidemia, and hypertension. Similar trends were observed for baseline characteristics stratified by quartiles of KDM-BA acceleration and PhenoAge acceleration.

**Table 1.** Baseline characteristics of study participants grouped by telomere length quartiles.

| Variables | Total | Telomere length |  |  |  | *p* value |
| --- | --- | --- | --- | --- | --- | --- |
|  |  | Q4 | Q3 | Q2 | Q1 |  |
| **N** | 311646 | 77912 | 77911 | 77911 | 77912 |  |
| **Age (years)** | 58.00 [50.00, 63.00] | 55.00 [47.00, 61.00] | 57.00 [49.00, 62.00] | 58.00 [51.00, 63.00] | 60.00 [53.00, 65.00] | <0.001 |
| **Women, n (%)** | 168249 (54.0) | 46842 (60.1) | 43183 (55.4) | 40641 (52.2) | 37583 (48.2) | <0.001 |
| **White, n (%)** | 294574 (94.5) | 72266 (92.8) | 73538 (94.4) | 74127 (95.1) | 74643 (95.8) | <0.001 |
| **TDI** | -2.20 [-3.67, 0.39] | -2.13 [-3.63, 0.51] | -2.19 [-3.67, 0.39] | -2.22 [-3.68, 0.33] | -2.24 [-3.68, 0.34] | <0.001 |
| **Education, n (%)** |  |  |  |  |  | <0.001 |
| College | 101091 (32.4) | 28303 (36.3) | 25913 (33.3) | 24385 (31.3) | 22490 (28.9) |  |
| High school | 35071 (11.3) | 9517 (12.2) | 8918 (11.4) | 8696 (11.2) | 7940 (10.2) |  |
| Middle school | 67420 (21.6) | 16538 (21.2) | 16993 (21.8) | 16990 (21.8) | 16899 (21.7) |  |
| Others | 108064 (34.7) | 23554 (30.2) | 26087 (33.5) | 27840 (35.7) | 30583 (39.3) |  |
| **BMI, Kg/m<sup>2</sup>** | 26.68 [24.11, 29.79] | 26.33 [23.81, 29.44] | 26.64 [24.06, 29.76] | 26.78 [24.20, 29.86] | 26.98 [24.39, 30.07] | <0.001 |
| **CRP, mg/L** | 1.30 [0.64, 2.67] | 1.19 [0.59, 2.50] | 1.27 [0.63, 2.61] | 1.32 [0.66, 2.70] | 1.41 [0.71, 2.85] | <0.001 |
| **Drinking status, n (%)** |  |  |  |  |  | <0.001 |
| Never | 13303 (4.3) | 3577 (4.6) | 3402 (4.4) | 3179 (4.1) | 3145 (4.0) |  |
| Previous | 10545 (3.4) | 2586 (3.3) | 2595 (3.3) | 2578 (3.3) | 2786 (3.6) |  |
| Current | 287103 (92.1) | 71568 (91.9) | 71750 (92.1) | 71978 (92.4) | 71807 (92.2) |  |
| **Smoking status, n (%)** |  |  |  |  |  | <0.001 |
| Never | 171217 (54.9) | 45181 (58.0) | 43698 (56.1) | 42177 (54.1) | 40161 (51.5) |  |
| Previous | 107241 (34.4) | 24874 (31.9) | 26162 (33.6) | 27418 (35.2) | 28787 (36.9) |  |
| Current | 31733 (10.2) | 7496 (9.6) | 7714 (9.9) | 7948 (10.2) | 8575 (11.0) |  |
| **Physical activity, n** |  |  |  |  |  | <0.001 |
| Regular | 122958 (39.5) | 31809 (40.8) | 31060 (39.9) | 30289 (38.9) | 29800 (38.2) |  |
| Excessive | 75514 (24.2) | 18426 (23.6) | 18780 (24.1) | 19133 (24.6) | 19175 (24.6) |  |
| Poor | 43693 (14.0) | 11063 (14.2) | 10928 (14.0) | 10956 (14.1) | 10746 (13.8) |  |
| **Sleep pattern, n (%)** |  |  |  |  |  | <0.001 |
| Healthy | 151320 (48.6) | 38635 (49.6) | 38006 (48.8) | 37521 (48.2) | 37158 (47.7) |  |
| Intermediate | 100598 (32.3) | 24512 (31.5) | 25132 (32.3) | 25319 (32.5) | 25635 (32.9) |  |
| Poor | 5807 (1.9) | 1337 (1.7) | 1439 (1.8) | 1475 (1.9) | 1556 (2.0) |  |
| **Diet pattern, n (%)** |  |  |  |  |  | <0.001 |
| Healthy | 117408 (37.7) | 30256 (38.8) | 29456 (37.8) | 29244 (37.5) | 28452 (36.5) |  |
| Intermediate | 145882 (46.8) | 35983 (46.2) | 36512 (46.9) | 36562 (46.9) | 36825 (47.3) |  |
| Poor | 35506 (11.4) | 8586 (11.0) | 8806 (11.3) | 8857 (11.4) | 9257 (11.9) |  |
| **Medical history, n (%)** |  |  |  |  |  |  |
| Hyperlipidemia | 144832 (46.5) | 33301 (42.7) | 35561 (45.6) | 37139 (47.7) | 38831 (49.8) | <0.001 |
| Hypertension | 170827 (54.8) | 38908 (49.9) | 41630 (53.4) | 43982 (56.5) | 46307 (59.4) | <0.001 |
| Diabetes mellitus | 17765 (5.7) | 3562 (4.6) | 4152 (5.3) | 4597 (5.9) | 5454 (7.0) | <0.001 |

Abbreviation: TDI, Townsend deprivation index; CRP, C-reactive protein; BMI, body mass index.

### Association between biological aging and the risk of AAA

During a median follow-up of 12.54 years (IQR: 11.87-13.16), a total of 1,339 (4.3‰) incident cases of AAA were reported. Overall, accelerated biological aging was significantly positively associated with an increased long-term risk of AAA (**Table 2**). Specifically, for each standard deviation decrease in TL, the risk of AAA increased by 20% (HR = 1.20, 95% CI = 1.14-1.27); for each standard deviation increase in KDM-BA acceleration, the risk increased by 21% (HR = 1.21, 95% CI = 1.12-1.29); and for each standard deviation increase in PhenoAge acceleration, the risk increased by 40% (HR = 1.40, 95% CI = 1.32-1.48). Consistently, compared with participants with longer TL, lower KDM-BA acceleration, or lower PhenoAge acceleration, those with shorter TL, higher KDM-BA acceleration, or higher PhenoAge acceleration had significantly higher risks of AAA. Furthermore, **Figure 1** demonstrated the potential dose-response relationships between biological aging and AAA risk. The results indicated a linear association between TL, KDM-BA acceleration, PhenoAge acceleration, and AAA occurrence (*p*-value for non-linearity > 0.05).

**Figure 1.** Association between biological aging and the risk of AAA using restricted cubic splines model with three knots. Abbreviation: AAA, abdominal aortic aneurysm; KDM-BA, Klemera-Doubal method biological age; PhenoAge, phenotypic age; HR, hazard ratio; CI, confidence interval.

**Table 2.**
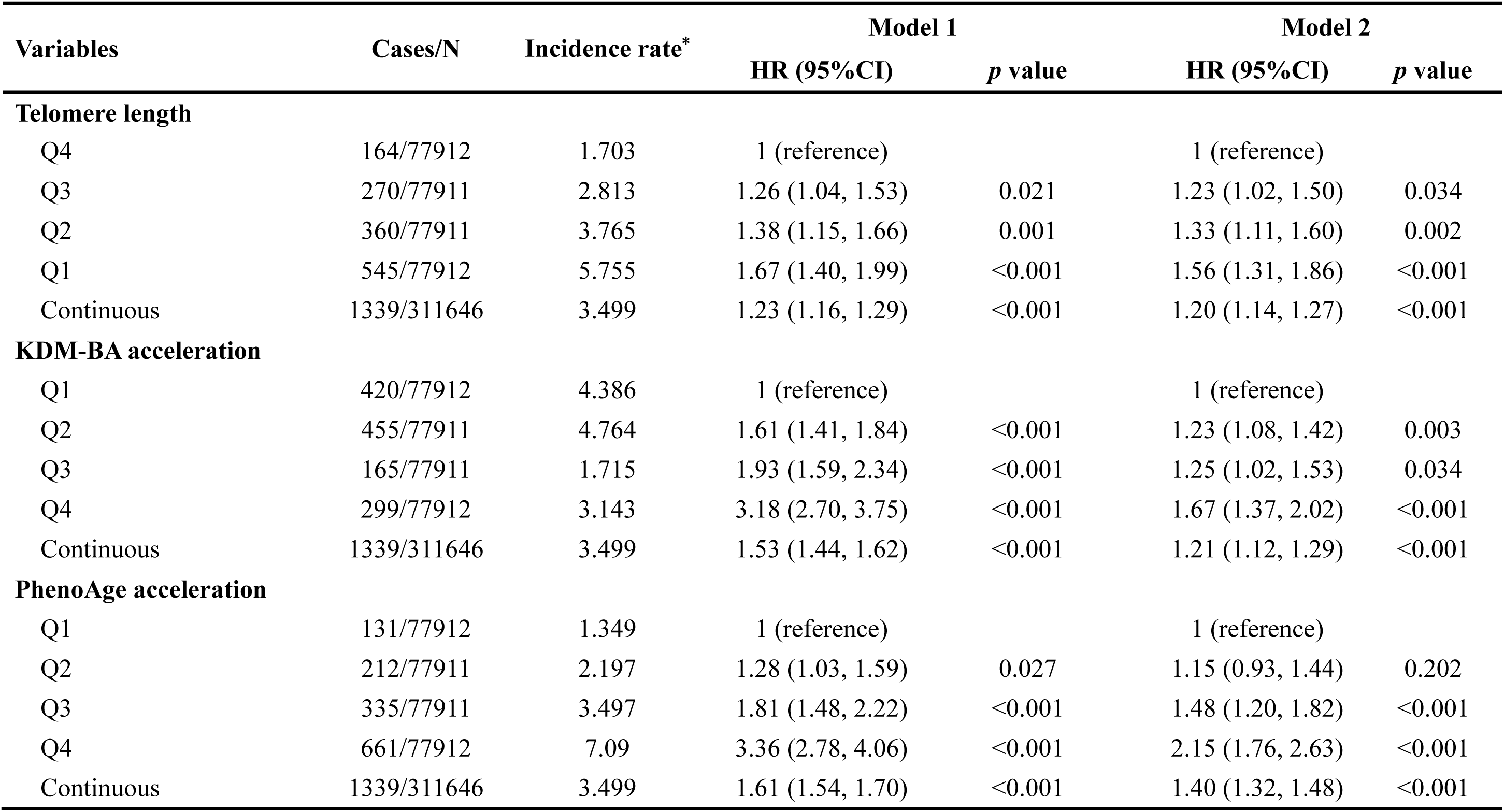
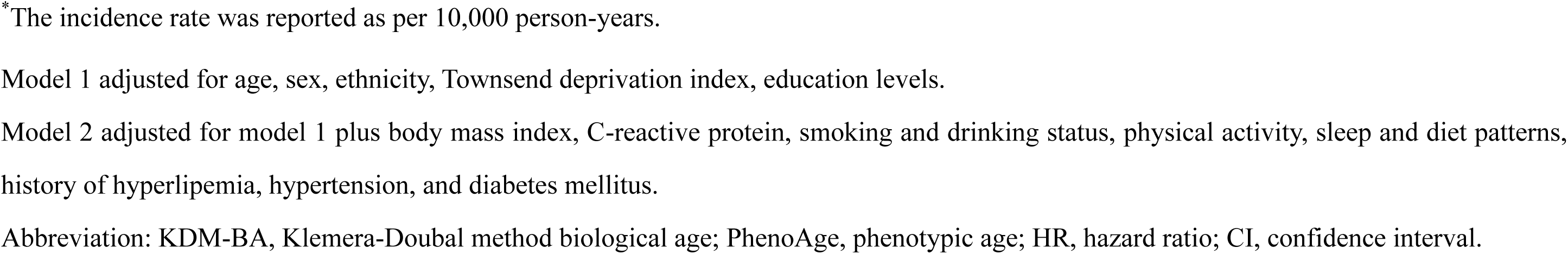
Association between biological aging and the risk of abdominal aortic aneurysm.

| Variables | Cases/N | Incidence rate* | Model 1 |  | Model 2 |  |
| --- | --- | --- | --- | --- | --- | --- |
|  |  |  | HR (95%CI) | *p* value | HR (95%CI) | *p* value |
| Telomere length |  |  |  |  |  |  |
| Q4 | 164/77912 | 1.703 | 1 (reference) |  | 1 (reference) |  |
| Q3 | 270/77911 | 2.813 | 1.26 (1.04, 1.53) | 0.021 | 1.23 (1.02, 1.50) | 0.034 |
| Q2 | 360/77911 | 3.765 | 1.38 (1.15, 1.66) | 0.001 | 1.33 (1.11, 1.60) | 0.002 |
| Q1 | 545/77912 | 5.755 | 1.67 (1.40, 1.99) | <0.001 | 1.56 (1.31, 1.86) | <0.001 |
| Continuous | 1339/311646 | 3.499 | 1.23 (1.16, 1.29) | <0.001 | 1.20 (1.14, 1.27) | <0.001 |
| KDM-BA acceleration |  |  |  |  |  |  |
| Q1 | 420/77912 | 4.386 | 1 (reference) |  | 1 (reference) |  |
| Q2 | 455/77911 | 4.764 | 1.61 (1.41, 1.84) | <0.001 | 1.23 (1.08, 1.42) | 0.003 |
| Q3 | 165/77911 | 1.715 | 1.93 (1.59, 2.34) | <0.001 | 1.25 (1.02, 1.53) | 0.034 |
| Q4 | 299/77912 | 3.143 | 3.18 (2.70, 3.75) | <0.001 | 1.67 (1.37, 2.02) | <0.001 |
| Continuous | 1339/311646 | 3.499 | 1.53 (1.44, 1.62) | <0.001 | 1.21 (1.12, 1.29) | <0.001 |
| PhenoAge acceleration |  |  |  |  |  |  |
| Q1 | 131/77912 | 1.349 | 1 (reference) |  | 1 (reference) |  |
| Q2 | 212/77911 | 2.197 | 1.28 (1.03, 1.59) | 0.027 | 1.15 (0.93, 1.44) | 0.202 |
| Q3 | 335/77911 | 3.497 | 1.81 (1.48, 2.22) | <0.001 | 1.48 (1.20, 1.82) | <0.001 |
| Q4 | 661/77912 | 7.09 | 3.36 (2.78, 4.06) | <0.001 | 2.15 (1.76, 2.63) | <0.001 |
| Continuous | 1339/311646 | 3.499 | 1.61 (1.54, 1.70) | <0.001 | 1.40 (1.32, 1.48) | <0.001 |

\*The incidence rate was reported as per 10,000 person-years.
Model 1 adjusted for age, sex, ethnicity, Townsend deprivation index, education levels.
Model 2 adjusted for model 1 plus body mass index, C-reactive protein, smoking and drinking status, physical activity, sleep and diet patterns, history of hyperlipemia, hypertension, and diabetes mellitus.
Abbreviation: KDM-BA, Klemera-Doubal method biological age; PhenoAge, phenotypic age; HR, hazard ratio; CI, confidence interval.

### Joint and interaction analysis of biological aging and genetic predisposition

The results in **Table S6** indicated that the constructed PRS score effectively predicted the risk of AAA occurrence. After further adjusting for PRS in Models 1 and 2, the significant association between biological aging indicators and AAA risk persisted (**Table S7**).

The joint effects of biological aging and genetic susceptibility on the risk of AAA were shown in **Figure 2A-C**. Overall, the risk of AAA increased with accelerated biological aging and higher levels of genetic susceptibility (*p*-value for trend < 0.001). Compared to participants with the lowest PRS and biological aging, those with the highest PRS and biological aging exhibited the highest risk of AAA (high PRS and Q1 of TL: HR = 1.81, 95% CI = 1.31-2.49; high PRS and Q4 of KDM-BA acceleration: HR = 2.43, 95% CI = 1.79-3.28; high PRS and Q4 of PhenoAge acceleration: HR = 3.27, 95% CI = 2.21-4.85). Additionally, at different levels of genetic susceptibility, higher levels of biological aging were positively associated with the occurrence of AAA (**Figure 2D-F**). No significant interaction was observed between genetic susceptibility and biological aging indicators regarding the risk of AAA (*p*-value for interaction > 0.05).

**Figure 2.** Separate and joint association of biological aging with the risk of AAA in individuals with different levels of genetic susceptibility. The result was presented as HR (95% CI) using the Cox proportional hazards model adjusted for age, sex, ethnicity, Townsend deprivation index, education levels, body mass index, C-reactive protein, smoking and drinking status, physical activity, sleep and diet patterns, history of hyperlipemia, hypertension, and diabetes mellitus Abbreviation: AAA, abdominal aortic aneurysm; KDM-BA, Klemera-Doubal method biological age; PhenoAge, phenotypic age; HR, hazard ratio; CI, confidence interval.

### Subgroup and sensitivity analyses

As shown in **Table 3**, subgroup analyses indicated that the association between shortened TL and the incidence of AAA was significant only in men, rather than in women (*p*-value for the interaction between sex and TL = 0.018). For other stratification factors, no significant differences were observed in the associations between biological aging indicators and AAA occurrence (*p*-value for the interaction > 0.05, **Table S8-9**). Sensitivity analyses, including the exclusion of participants who reported AAA in the first year (**Table S10**) and the application of Fine-Gray competing risk regression models (**Figure S3**), consistently supported the strong association between high levels of biological aging and an increased incidence of AAA.

**Table 3.** Subgroup analysis for the association between telomere length with the risk of abdominal aortic aneurysm.

| Subgroup | Cases/N | Telomere length |  |  |  | *p* for interaction |
| --- | --- | --- | --- | --- | --- | --- |
|  |  | Q4 | Q3 | Q2 | Q1 |  |
| **Age** |  |  |  |  |  | 0.409 |
| <65 | 665/253654 | 1 (reference) | 1.31 (1.01, 1.71) | 1.63 (1.27, 2.10) | 1.94 (1.52, 2.46) |  |
| ≥65 | 674/57992 | 1 (reference) | 1.06 (0.83, 1.36) | 1.26 (1.00, 1.60) | 1.63 (1.30, 2.03) |  |
| **Sex** |  |  |  |  |  | 0.018 |
| Women | 205/168249 | 1 (reference) | 0.84 (0.55, 1.29) | 0.79 (0.52, 1.19) | 1.00 (0.68, 1.47) |  |
| Men | 1134/143397 | 1 (reference) | 1.32 (1.07, 1.62) | 1.45 (1.19, 1.76) | 1.69 (1.40, 2.04) |  |
| **Ethnicity** |  |  |  |  |  | 0.829 |
| White | 1308/294574 | 1 (reference) | 1.26 (1.04, 1.53) | 1.32 (1.10, 1.60) | 1.56 (1.31, 1.86) |  |
| Non-White | 31/17072 | 1 (reference) | 1.21 (0.29, 5.09) | 1.35 (0.35, 5.16) | 1.99 (0.56, 7.03) |  |
| **TDI** |  |  |  |  |  | 0.557 |
| Low | 411/104004 | 1 (reference) | 1.55 (1.08, 2.24) | 1.64 (1.15, 2.33) | 1.84 (1.31, 2.58) |  |
| Moderate | 445/103925 | 1 (reference) | 1.05 (0.77, 1.44) | 1.02 (0.75, 1.38) | 1.29 (0.97, 1.71) |  |
| High | 483/103717 | 1 (reference) | 1.19 (0.85, 1.66) | 1.47 (1.07, 2.02) | 1.65 (1.22, 2.24) |  |
| **Education** |  |  |  |  |  | 0.519 |
| College | 223/101091 | 1 (reference) | 1.62 (1.01, 2.61) | 1.43 (0.89, 2.29) | 1.83 (1.17, 2.86) |  |
| High school | 109/35071 | 1 (reference) | 2.06 (0.99, 4.31) | 1.99 (0.97, 4.09) | 2.39 (1.19, 4.76) |  |
| Middle school | 289/67420 | 1 (reference) | 1.12 (0.75, 1.69) | 1.28 (0.87, 1.88) | 1.26 (0.87, 1.83) |  |
| Others | 718/108064 | 1 (reference) | 1.15 (0.89, 1.49) | 1.22 (0.96, 1.57) | 1.61 (1.27, 2.02) |  |
| Hypertension |  |  |  |  |  | 0.647 |
| No | 235/140819 | 1 (reference) | 1.82 (1.08, 3.06) | 1.82 (1.10, 3.01) | 2.12 (1.30, 3.44) |  |
| Yes | 1104/170827 | 1 (reference) | 1.18 (0.96, 1.44) | 1.26 (1.04, 1.53) | 1.50 (1.24, 1.80) |  |
| Hyperlipidemia |  |  |  |  |  | 0.266 |
| No | 344/166814 | 1 (reference) | 1.47 (0.97, 2.23) | 1.72 (1.16, 2.56) | 1.81 (1.23, 2.65) |  |
| Yes | 995/144832 | 1 (reference) | 1.09 (0.88, 1.34) | 1.21 (0.99, 1.48) | 1.45 (1.19, 1.75) |  |
| Diabetes |  |  |  |  |  | 0.516 |
| No | 1201/293881 | 1 (reference) | 1.20 (0.97, 1.47) | 1.34 (1.10, 1.63) | 1.55 (1.29, 1.87) |  |
| Yes | 138/17765 | 1 (reference) | 1.25 (0.73, 2.14) | 1.15 (0.68, 1.96) | 1.32 (0.79, 2.21) |  |

Adjusted for age, sex, ethnicity, TDI, education levels body mass index, C-reactive protein, smoking and drinking status, physical activity, sleep and diet patterns, history of hyperlipemia, hypertension, and diabetes mellitus. Abbreviation: TDI, Townsend deprivation index.

## Discussion

In this study, we utilized the large-scale prospective cohort of the UK Biobank to investigate the association between clinical biomarker-based biological aging and the long-term risk of AAA. The main findings are as follows: accelerated biological aging (reflected by shortened TL, higher KDM-BA, and PhenoAge acceleration) was significantly associated with an increased risk of AAA. This association was independent of genetic risk assessed by PRS and demonstrated a joint effect with genetic predisposition. Furthermore, sex appeared to modify the relationship between TL and AAA risk, with the association between shortened TL and an increased risk of AAA being significant only in men.

The association between biological aging and cardiovascular diseases has gradually been recognized[18, 19], and accurately measuring biological age is key to further studying this relationship[20]. To this end, various indicators for assessing an individual’s biological age have been proposed, each of which is distinctive in reflecting different dimensions and mechanisms of the aging process. Telomeres are tandemly repeated nucleotide sequences located at the ends of chromosomes, and TL is considered a key indicator of biological aging[21]. When telomeres shorten to a critical threshold, an irreparable DNA damage response is triggered, leading to the cessation of cell division and, consequently, aging[22]. A study by Atturu et al.[23] showed that, compared to the control group, patients with AAA had shorter leukocyte telomeres, and this telomere shortening was significantly associated with AAA (odds ratio [OR] = 2.30, 95% CI =1.28-4.13). However, this was a case-control study with a limited sample size (N = 373), and it did not establish a longitudinal association between TL and the occurrence of AAA. This study partially addressed this issue and provided stronger evidence. We also found an interesting interaction between TL and sex. The association between biological aging and AAA risk was only significant in men. The remodeling of the extracellular matrix has been found to be more pronounced in the aging process of men compared to women, as evidenced by increased aortic diameter and stiffness[24]. This may be attributed to the protective effects of estrogen[25]. In addition, several composite biomarker algorithms, such as KDM-BA and PhenoAge, have been proposed in recent years to estimate an individual’s biological age. Compared to single indicators like TL, these algorithms are more reliable and comprehensive, as they integrate a range of clinical biomarkers and anthropometric data while also considering multiple dimensions, such as metabolism, immunity, inflammation, and organ homeostasis[26]. Furthermore, their high accessibility and affordability further enhance their potential for widespread application.

Biological aging may contribute to the development of AAA through various mechanisms. Chronic low-grade inflammation is a significant characteristic of aging and a key mechanism in AAA development. Senescent cells secrete a complex array of factors, including pro-inflammatory cytokines, chemokines, growth factors, and matrix metalloproteinases[27]. These factors can recruit various inflammatory cells to infiltrate the vascular wall, creating an inflammatory microenvironment that compromises aortic structural integrity and promotes the occurrence of AAA[28]. Additionally, oxidative stress may partly explain the association between biological aging and AAA. Research has shown that aging is related to the dysfunction of nuclear factor E2-related factor 2 in the aorta, exacerbating oxidative stress and increasing sensitivity to reactive oxygen species-mediated damage[29]. Moreover, the phenotypic transformation of smooth muscle cells is another important mechanism in the development of aortic aneurysms. Under pressure induction, aging vascular smooth muscle cells secrete fibroblast growth factor 9, causing adjacent normal vascular smooth muscle cells to undergo phenotypic transformation, which may contribute to the formation of AAA[30].

The strength of this study lies in establishing, for the first time, a longitudinal association between biological aging and the long-term risk of AAA. The large sample size and high-quality data from the UK Biobank prospective cohort ensured the credibility of the study results. Additionally, this study is notably comprehensive, utilizing three different indicators to assess individual biological aging levels: TL, a key marker of aging, and KDM-BA and PhenoAge, which are calculated based on multidimensional biomarkers. Furthermore, we also took into account the role of genetic susceptibility in this association. This study also has some limitations. First, as an observational study, it cannot establish causality or clarify the specific mechanisms underlying the association between biological aging and AAA. Second, biological aging was assessed solely through baseline data, preventing further exploration of the relationship between longitudinal changes in biological aging and AAA risk. Although multiple confounding factors were adjusted for, the study cannot rule out the potential influence of important confounders not available in the UK Biobank. Additionally, since the participants of the UK Biobank were primarily White, the generalizability of the findings to other populations needs further confirmation.

## Conclusion

Biomarker-based biological aging is significantly associated with the long-term risk of AAA, independent of genetic susceptibility. These findings highlight the potential utility of biological aging assessments in identifying high-risk populations for AAA, providing valuable insights for its prevention and management.

## Data Availability

The data supporting the findings of this study were obtained from the UK Biobank Resource under Application No. 107175. These data can be accessed by submitting an application through the UK Biobank official website (www.ukbiobank.ac.uk).

## Declarations

### Ethics approval and consent to participate

This study was approved by the North West Multi-Centre Research Ethics Committee (Reference: 11/NW/0382). All participants provided written informed consent prior to participation.

### Consent for publication

Not applicable.

### Author contributions

This study was conceived and designed by Yiyang Tang. Xinyi Zhou and Zaixin Yu performed the data analysis and drafted the manuscript. Mukamengjiang Juaiti and Lihuang Zha contributed to the statistical analysis. Baohua Peng assisted with data interpretation and visualization. Chengshuang Xiao revised the manuscript and provided constructive feedback. All authors reviewed and approved the final manuscript.

## Acknowledgements

The authors sincerely thank the management team and participants of the UK Biobank for their invaluable contributions to this study.

